# Real-world effectiveness of early molnupiravir and nirmatrelvir/ritonavir among hospitalized, non-oxygen-dependent COVID-19 patients on admission during Hong Kong’s Omicron BA.2 wave: an observational study

**DOI:** 10.1101/2022.05.19.22275291

**Authors:** Carlos K.H. Wong, Ivan C.H. Au, Kristy T.K. Lau, Eric H.Y. Lau, Benjamin J. Cowling, Gabriel M. Leung

## Abstract

**Background:** Effectiveness of oral antivirals in mild-to-moderate COVID-19 patients is urgently needed. This retrospective cohort study aims to evaluate the clinical and virologic outcomes associated with molnupiravir and nirmatrelvir/ritonavir use in COVID-19 patients during a pandemic wave dominated by the Omicron BA.2 subvariant.

**Methods:** We analyzed data from a territory-wide retrospective cohort of hospitalized patients with confirmed diagnosis of SARS-CoV-2 infection from 26th February 2022 to 26th April 2022 in Hong Kong. Oral antiviral users were matched with controls using propensity-score matching in a ratio of 1:1. Study outcomes were all-cause mortality, a composite outcome of disease progression (all-cause mortality, initiation of invasive mechanical ventilation [IMV], intensive care unit admission, or the need for oxygen therapy) and their individual outcomes, and time to achieving lower viral burden of cycle threshold (Ct) value ≥30 cycles. Hazard ratios (HR) of event outcomes were estimated using Cox regression models.

**Results:** Among 40,776 hospitalized patients with SARS-CoV-2 infection over a mean follow-up of 41.3 days with 925,713 person-days, this study included 1,856 molnupiravir users, 890 nirmatrelvir/ritonavir users and 2,746 control patients not initially requiring oxygen therapy at baseline after propensity-score matching. Oral antiviral use was associated with significantly lower risks of all-cause mortality (molnupiravir: HR=0.48, 95%CI=0.40-0.59, p<0.0001; nirmatrelvir/ritonavir: HR=0.34, 95%CI=0.23-0.50, p<0.0001), the composite outcome of disease progression (molnupiravir: HR=0.60, 95%CI=0.52-0.69, p<0.0001; nirmatrelvir/ritonavir: HR=0.57, 95%CI=0.45-0.72, p<0.0001), and the need for oxygen therapy (molnupiravir: HR=0.69, 95%CI=0.57-0.83, p=0.00011; nirmatrelvir/ritonavir: HR=0.73, 95%CI=0.54-0.97, p=0.032) than non-use. Time to achieving lower viral burden was significantly shorter among oral antiviral users than matched controls (molnupiravir: HR=1.38, 95%CI=1.15-1.64, p=0.0046; nirmatrelvir/ritonavir: HR=1.38, 95%CI=1.07-1.78, p=0.013).

**Conclusions:** Against Omicron BA.2, initiation of novel oral antiviral treatment in hospitalized patients not requiring any oxygen therapy was associated with lower risks of all-cause mortality and disease progression, in addition to achieving low viral burden faster. Our findings support the early use of oral antivirals in COVID-19 patients who do not require supplemental oxygen on admission.

**Funding:** Health and Medical Research Fund, Food and Health Bureau, Government of the Hong Kong SAR

**Research in context:** *Evidence before this study:* The medical and research community are actively exploring the use of oral antivirals in COVID-19 patients to lower their risks of hospitalization and death, and to reduce the burden on healthcare systems. We searched Scopus and PubMed for studies until 13th May 2022 using the search terms “SARS-CoV-2 OR COVID-19” AND “molnupiravir OR Lagevrio OR EIDD-2801” OR “nirmatrelvir OR Paxlovid OR PF-07321332”. Major studies examining the safety and efficacy of molnupiravir include MOVe-IN and MOVe-OUT trials conducted in hospitalized and non-hospitalized COVID-19 patients, respectively. Clinical evidence for the use of ritonavir-boosted nirmatrelvir came from the EPIC-HR trial conducted among non-hospitalized adults with COVID-19. While no clinical benefits have been observed with molnupiravir use in the inpatient setting among patients with moderate-to-severe COVID-19, early initiation of molnupiravir or nirmatrelvir/ritonavir within 5 days of symptom onset in non-hospitalized patients with mild-to-moderate COVID-19 and risk factors for progression to severe disease has been associated with relative risk reduction of hospitalization or death by 30% and 88%, respectively. Notably, these clinical trials were conducted prior to the prevalence of Omicron variant, and the efficacy of oral antivirals against this current variant of concern can only be inferred from experimental evidence to date. Real-world evidence of oral antiviral use in patients with SARS-CoV-2 infection of Omicron variant is lacking.

*Added value of this study:* To the best of our knowledge, this is the first real-world study exploring the inpatient use of oral antivirals during a pandemic wave dominated by SARS-CoV-2 Omicron variant. We conducted a territory-wide, retrospective cohort study to examine the effectiveness of molnupiravir and nirmatrelvir/ritonavir in COVID-19 patients who did not require supplemental oxygen on admission in Hong Kong. Early initiation of oral antivirals within 2 days of admission was associated with significantly lower risks of all-cause mortality and disease progression, in addition to achieving low viral burden faster than their respective matched controls. Oral antiviral use was also associated with a reduced need for oxygen therapy than non-use.

*Implications of all the available evidence:* Current guidelines are now prioritizing the distribution of oral antivirals to those who do not require supplemental oxygen, but who are at the highest risk of disease progression. Our study cohort reflected such prescription pattern in real-world clinical practice, consisting of mostly the elderly with multiple pre-existing comorbidities and who had not been fully vaccinated. The antiviral effect and mortality benefit observed in this patient cohort support the use of oral antivirals in COVID-19 patients who do not require supplemental oxygen on admission during a pandemic wave of Omicron variant. Ongoing research will inform the safety and effectiveness of oral antivirals in specific patient populations (by vaccination status and viral variants), drug combinations, and different healthcare settings.

## Introduction

In the midst of the coronavirus disease 2019 (COVID-19) pandemic, various drugs have been repurposed or developed for treating patients with SARS-CoV-2 infection. In December 2021, molnupiravir (Lagevrio) and ritonavir-boosted nirmatrelvir (Paxlovid) are two oral antivirals that have been granted Emergency Use Authorization (EUA) by the U.S. Food and Drug Administration (FDA) for the treatment of non-hospitalized patients with mild-to-moderate COVID-19, who are at risk of progression to severe disease, so as to reduce the burden on healthcare systems by lowering their risk of hospitalization or death.^1,2^

While both molnupiravir and nirmatrelvir/ritonavir are indicated for non-hospitalized patients with mild-to-moderate COVID-19 within 5 days of symptom onset should they be at risk of progression to severe disease, current guidelines give priority to nirmatrelvir/ritonavir (relative risk reduction by 88%) and another antiviral remdesivir (by 87%) that have demonstrated higher efficacy than molnupiravir (by 30%) in reducing hospitalization or death among COVID-19 patients not requiring hospitalization or supplemental oxygen.^1-4^ Notably, several concerns and research gaps remain in the use of the two oral antivirals, for instance, if initiation in asymptomatic COVID-19 patients is appropriate, the lack of clinical data in treating patients infected with specific VOC, and their safety and efficacy in vaccinated individuals with breakthrough infections.^5-7^ Furthermore, efficacy of molnupiravir as illustrated in the MOVe-OUT trial has been questioned in light of its premature termination, imbalances in risk factors and COVID-19 severity of patients at baseline, results with borderline statistical significance and of uncertain clinical significance, and discrepancies between interim and full analyses that could not be fully explained by differences in patient characteristics.^8-10^

Real-world evidence on the effectiveness of molnupiravir and nirmatrelvir/ritonavir in COVID-19 patients is urgently needed.^11^ This retrospective cohort study aims to evaluate the clinical and virologic outcomes associated with molnupiravir and nirmatrelvir/ritonavir use in COVID-19 patients during a community epidemic dominated by the Omicron BA.2 subvariant. While both oral antivirals are now indicated for non-hospitalized COVID-19 patients who are at high risk of disease progression, the current analysis focuses on their effectiveness in hospitalized patients with COVID-19 who do not initially require any oxygen therapy on admission.

## Methods

### Study Design

A territory-wide, retrospective cohort study was used to examine the effectiveness of oral antivirals (molnupiravir or nirmatrelvir/ritonavir) in hospitalized adult patients with COVID-19 without oxygen therapy in the Hong Kong Special Administrative Region, China, during the observation period from 26th February 2022 to 5th May 2022.

### Data Source and Study Population

Electronic health records of patients with COVID-19 were retrieved from the Hospital Authority (HA), a statutory provider of public inpatient services and primary public outpatient services in Hong Kong. Electronic health records included demographics, date of registered death, hospital admission, emergency department visits, diagnoses, prescription and drug dispensing records, procedures, and laboratory tests. The HA linked the health records and anonymized population-based vaccination records of individuals provided by the Department of Health using unique identification numbers (Hong Kong Identity Card or foreign passport number). The database has been widely used for high-quality studies to evaluate the safety and effectiveness of drug treatments for COVID-19 at a population level.^12,13^ For all-cause mortality, data were extracted and ascertained from the Hong Kong Death Registry, which would allow us to capture any death events of patients occurring beyond hospital discharge (outside the hospital setting). Our cohort comprised patients with positive results of reverse transcription-polymerase chain reaction (RT-PCR) or rapid antigen test who were admitted to isolation wards at local public hospitals from 26th February 2022 to 26th April 2022. Patients were eligible for inclusion if they had been admitted within 3 days of their COVID-19 diagnosis date, or if COVID-19 diagnosis was confirmed within 3 days of their admission date, so as to account for any potential time lag in the confirmation of cases during an upsurge of patients with SARS-CoV-2 infection. The index date was defined as the date of hospital admission (day 0). Patients who were admitted to hospital with COVID-19 before 26th February 2022 (the date when molnupiravir first became locally available), after 26th April 2022 (less than 1 week of follow-up), or beyond 5 days of symptom onset, aged <18 years, or with oxygen support or mechanical ventilation on the index date, were excluded. Patients with drug contraindications to nirmatrelvir/ritonavir (i.e. use of amiodarone, apalutamide, lumacaftor/ivacaftor, ivosidenib, rifampicin, rifapentine, carbamazepine, St John’s Wort, primidone, phenobarbital, or phenytoin in the past 6 months prior to the baseline),^14^ severe renal impairment^2^ (eGFR <30mL/min/1.73m^2^, dialysis, or renal transplantation), or severe liver impairment^2^ (cirrhosis, hepatocellular carcinoma, or liver transplantation) at baseline were excluded from the current analysis to further mitigate confounding by indication as much as possible, and restrict the sample to those who were as equally eligible to receive either molnupiravir or nirmatrelvir/ritonavir treatment as possible.

This study was approved by the institutional review board of the University of Hong Kong / Hospital Authority Hong Kong West Cluster (reference no. UW 20-493). Given the extraordinary nature of the COVID-19 pandemic, individual patient-informed consent was not required for this retrospective cohort study using anonymized data.

### Treatment Exposure and Follow-up Period

Hospitalized patients with COVID-19 without oxygen therapy and receiving early (i) molnupiravir or (ii) nirmatrelvir/ritonavir treatment at public hospitals during the observation period were defined as (i) molnupiravir users and (ii) nirmatrelvir/ritonavir users, respectively. As all public hospitals in Hong Kong are managed by the Hospital Authority, oral antivirals were prescribed to COVID-19 patients as clinically appropriate based on the same set of standard treatment protocols, and both oral antivirals were equally accessible across all public hospitals during the study period (since 26th February 2022 for molnupiravir,^15^ and nirmatrelvir/ritonavir was locally available since 16th March 2022^16^). We defined treatment exposure period at 2 days within admission to mitigate potential immortal time bias between treatment initiation and admission.^17-20^ Controls were selected from the cohort of hospitalized patients with COVID-19 without oxygen therapy who did not receive oral antivirals (molnupiravir and/or nirmatrelvir/ritonavir) during the observation period, using the propensity-score in a ratio of 1:1, and considering the time period of admission. Patients were observed from the index date until registered death, the occurrence of outcome events, crossover of oral antiviral treatment, or the end of the observation period (5th May 2022), whichever came first.

### Outcomes

The primary outcome of this study was all-cause mortality. Secondary outcomes were a composite outcome of disease progression (all-cause mortality, initiation of invasive mechanical ventilation [IMV], intensive care unit [ICU] admission, or the need for oxygen therapy) and their individual outcomes, and time to achieving lower viral burden of cycle threshold (Ct) value ≥30 cycles. Viral burden information at baseline might not be immediately available for a minority of patients who were admitted based on positive rapid antigen test; and quantitative viral burden was not assessed as a routine procedure, especially during the peak of Omicron BA.2 epidemic when public hospitals were overwhelmed with cases. Hospital length of stay (LOS) was also determined for discharged survivors. In response to an upsurge of COVID-19 cases during the study period and the limited number of hospital beds, the HA had revised their discharge criteria on 26th February 2022 to allow patients hospitalized with COVID-19 to be discharged as soon as they were deemed clinically stable by their attending physicians, and provided that their residential premises were suitable for isolation or they would be accepted by community isolation facilities, where they would continue their isolation until negative test results were obtained (on days 6 and 7 for fully vaccinated individuals [with at least two doses]; and day 14 for those not fully vaccinated [unvaccinated or vaccinated with only one dose]).^21^

Over the follow-up period, changes in the proportion of patients with the respective clinical status (namely in-hospital death, on invasive mechanical ventilation, without invasive mechanical ventilation, and discharged) were compared between oral antiviral and respective control groups.

### Baseline Covariates

Baseline covariates of patients included age, sex, regions, nursing home residents, symptom onset date reported, date of hospital admission, nosocomial infection (defined as hospitalization before COVID-19 diagnosis), time period of hospital admission, Charlson Comorbidity Index (CCI), any previous SARS-CoV-2 infection (defined as a recorded medical history of confirmed SARS-CoV-2 infection), COVID-19 vaccination status (fully vaccinated as having at least two doses of Comirnaty or three doses of CoronaVac), concomitant treatments initiated on the index date (antibiotics, dexamethasone and other systemic steroids, interferon-β-1b, baricitinib, and tocilizumab), and laboratory parameters on admission (Ct value, lactate dehydrogenase, C-reactive protein, and lymphocyte count).

### Statistical Analyses

Propensity-score models conditional on the aforementioned baseline covariates without first-order interactions in a logistic regression model was performed, and the propensity of receiving each oral antiviral was estimated in an approach of caliper matching without replacement, with a caliper width of 0.05. Missing laboratory parameters (Supplementary Table 1, Appendix p.2) for oral antiviral users were imputed 20 times using other parameters in the propensity-score model.^22^ We applied Rubin’s rules to pool the treatment effects estimated from the 20 independent imputed datasets.^23^ We used the standardized mean difference (SMD) to assess the balance of each baseline covariate between the groups before and after propensity-score matching, with SMD greater than 0.1 indicating covariate imbalance.^24^

Hazard ratios (HR) with 95% confidence intervals (CI) of each outcome between oral antiviral users and non-users were estimated using Cox regression models. Since there is no evidence that the proportional-hazards assumption has been violated using Schoenfeld residuals, we assumed the proportionality of the hazard ratios in primary analysis. A cluster-robust sandwich variance-covariance estimator was used in all the Cox regression models to account for the correlation within the propensity-score match. Analyses were conducted among the following patient subgroups: age (≤65 or □65 years), fully vaccinated or not, region, study period (before or after 16th March 2022, the date since both oral antivirals were available across public hospitals), and with and without symptom onset date reported. Sensitivity analyses were performed by 1) including only patients with complete 28-day follow-up (i.e. inclusion period from 26th February to 7th April 2022), and 2) using the observed baseline characteristics without laboratory parameters (without multiple imputation) for propensity-score model. We re-matched baseline covariates, and constructed a new propensity score model for each subgroup and sensitivity analysis.

All statistical analyses were performed using Stata version 17 (StataCorp LP, College Station, TX). All significance tests were 2-tailed, where P-value <0.05 was considered statistically significant.

### Role of the funder

The funder had no role in the data collection, analysis, interpretation, writing of the manuscript, and the decision to submit.

## Results

In this territory-wide, retrospective cohort study, a total of 40,776 hospitalized patients with confirmed diagnosis of SARS-CoV-2 infection over a mean follow-up of 41.3 days (standard deviation: 18.7) with 925,713 person-days were identified from 26th February 2022 to 26th April 2022. This study included 1,856 molnupiravir users, 890 nirmatrelvir/ritonavir users and 2,746 control patients not initially requiring oxygen therapy at baseline after propensity-score matching. (Figure 1). Baseline characteristics of oral antiviral and control groups before matching are presented in Table 1. After 1:1 propensity-score matching, this analysis included 1,856 molnupiravir users (with 1,856 matched controls) and 890 nirmatrelvir/ritonavir users (with 890 matched controls) with COVID-19 not initially requiring any oxygen therapy at baseline. After matching, propensity score distribution of oral antiviral and matched control groups were highly overlapping (Supplementary Figure 1, Appendix p.13) while baseline characteristics of patients were balanced between oral antiviral and matched control groups with all SMDs ≤0.1 (Table 1 and Supplementary Table 2, Appendix p.3). The median duration from symptom onset to molnupiravir initiation was 1 (interquartile range [IQR]: 1-3) day, and that from symptom onset to nirmatrelvir/ritonavir initiation was 1 (IQR: 1-3) day. The proportion of molnupiravir who received molnupiravir 800mg twice daily for 5 days was 96.7% (1,795 of 1,856), while the proportion of nirmatrelvir/ritonavir users who completed the 5-days regimen (nirmatrelvir 300mg with ritonavir 100mg twice daily for 5 days) was 98.8% (880 of 890).

**Figure 1.**
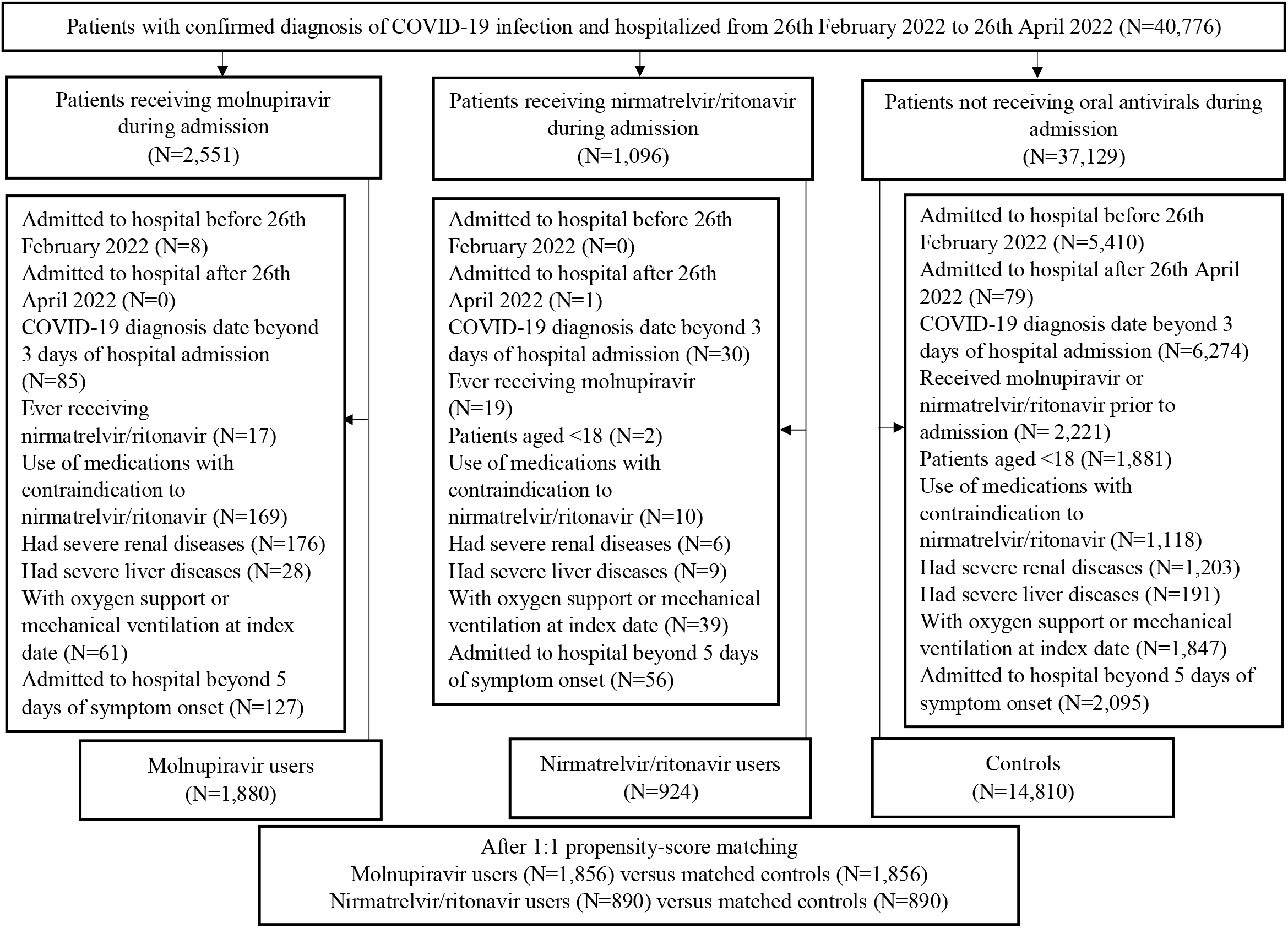
Identification of molnupiravir users, nirmatrelvir/ritonavir users, and their matched controls among patients hospitalized with COVID-19 not requiring oxygen therapy from 26th February 2022 to 26th April 2022 in Hong Kong

**Table 1.** Baseline characteristics of inpatients with COVID-19 in (a) molnupiravir and respective matched control groups, and (b) nirmatrelvir-ritonavir and respective matched control groups before 1:1 propensity score matching

| Baseline characteristics | Before 1:1 propensity score matching |  |  |  |  |  | After 1:1 propensity score matching |  |  |  |
| --- | --- | --- | --- | --- | --- | --- | --- | --- | --- | --- |
|  | Molnupiravir<br>(n=1,880) |  | Nirmatrelvir/<br>ritonavir<br>(n=924) |  | Control<br>(n=14,810) |  | Molnupiravir<br>vs Control | Nirmatrelvir/<br>ritonavir vs<br>Control | Molnupiravir<br>vs Control | Nirmatrelvir/<br>ritonavir vs<br>Control |
|  | N / Mean | % / SD | N / Mean | % / SD | N / Mean | % / SD | SMD | SMD | SMD | SMD |
| Age, years† | 80.8 | 13.0 | 77.2 | 14.1 | 74.3 | 18.7 | 0.36 | 0.16 | 0.04 | 0.05 |
| 18-40 | 27 | (1.4%) | 29 | (3.1%) | 1,313 | (8.9%) |  |  |  |  |
| 40-65 | 207 | (11.0%) | 132 | (14.3%) | 2,474 | (16.7%) | 0.40 | 0.26 | 0.10 | 0.07 |
| >65 | 1,646 | (87.6%) | 763 | (82.6%) | 11,023 | (74.4%) |  |  |  |  |
| Sex |  |  |  |  |  |  |  |  |  |  |
| Male | 925 | (49.2%) | 462 | (50.0%) | 7,500 | (50.6%) | 0.03 | 0.01 | 0.02 | 0.02 |
| Female | 955 | (50.8%) | 462 | (50.0%) | 7,310 | (49.4%) |  |  |  |  |
| Regions |  |  |  |  |  |  |  |  |  |  |
| Hong Kong Island | 502 | (26.7%) | 187 | (20.2%) | 2,297 | (15.5%) | 0.29 | 0.13 | 0.05 | 0.07 |
| Kowloon | 607 | (32.3%) | 288 | (31.2%) | 4,978 | (33.6%) |  |  |  |  |
| New Territories | 770 | (41.0%) | 446 | (48.3%) | 7,511 | (50.7%) |  |  |  |  |
| Others | 1 | (0.1%) | 3 | (0.3%) | 24 | (0.2%) |  |  |  |  |
| Nursing home residents | 579 | (30.8%) | 130 | (14.1%) | 5,003 | (33.8%) | 0.06 | 0.47 | 0.03 | 0.01 |
| Symptom onset date reported | 1,008 | (53.6%) | 404 | (43.7%) | 5,786 | (39.1%) | 0.29 | 0.09 | 0.01 | 0.03 |
| Time from symptom onset to hospitalization, days |  |  |  |  |  |  |  |  |  |  |
| 0 | 394 | (39.1%) | 160 | (39.6%) | 2,050 | (35.4%) | 0.08 | 0.09 | 0.01 | 0.01 |
| 1-5 | 614 | (60.9%) | 244 | (60.4%) | 3,736 | (64.6%) |  |  |  |  |
| Nosocomial infection | 45 | (2.4%) | 30 | (3.2%) | 968 | (6.5%) | 0.20 | 0.15 | 0.02 | 0.02 |
| Time of admission |  |  |  |  |  |  |  |  |  |  |
| February/March 2022 | 1,588 | (84.5%) | 626 | (67.8%) | 12,963 | (87.5%) | 0.09 | 0.13 | 0.06 | 0.01 |
| April 2022 | 292 | (15.5%) | 298 | (32.3%) | 1,847 | (12.5%) |  |  |  |  |
| Charlson's Index† | 5.8 | 1.9 | 5.1 | 1.7 | 5.0 | 2.4 | 0.33 | 0.03 | 0.01 | 0.02 |
| 1-4 | 459 | (24.4%) | 312 | (33.8%) | 5,312 | (35.9%) |  |  |  |  |
| 5-6 | 878 | (46.7%) | 465 | (50.3%) | 5,951 | (40.2%) | 0.25 | 0.24 | 0.06 | 0.09 |
| 7-14 | 543 | (28.9%) | 147 | (15.9%) | 3,547 | (24.0%) |  |  |  |  |
| Previous SARS-CoV-2 infection | 0 | (0.0%) | 0 | (0.0%) | 3 | (0.0%) | 0.02 | 0.02 | NA | NA |
| Fully vaccinated* | 116 | (6.2%) | 97 | (10.5%) | 1,328 | (9.0%) | 0.11 | 0.05 | 0.00 | 0.03 |
| Concomitant treatments initiated at admission |  |  |  |  |  |  |  |  |  |  |
| Antibiotics | 222 | (11.8%) | 158 | (17.1%) | 1,785 | (12.1%) | 0.01 | 0.14 | 0.00 | 0.00 |
| Immunomodulators | 260 | (13.8%) | 106 | (11.5%) | 3,258 | (22.0%) | 0.21 | 0.28 | 0.04 | 0.01 |
| Dexamethasone | 240 | (12.8%) | 87 | (9.4%) | 2,989 | (20.2%) | 0.20 | 0.31 | 0.04 | 0.00 |
| Other systemic steroid | 18 | (1.0%) | 22 | (2.4%) | 244 | (1.6%) | 0.06 | 0.05 | 0.00 | 0.04 |
| Interferon-β-1b | 10 | (0.5%) | 4 | (0.4%) | 195 | (1.3%) | 0.08 | 0.10 | 0.04 | 0.00 |
| Baricitinib | 0 | (0.0%) | 4 | (0.4%) | 29 | (0.2%) | 0.06 | 0.04 | 0.06 | 0.02 |
| Tocilizumab | 0 | (0.0%) | 0 | (0.0%) | 8 | (0.1%) | 0.03 | 0.03 | 0.03 | NA |
| Laboratory parameters at admission† |  |  |  |  |  |  |  |  |  |  |
| Cycle threshold value, cycle | 22.3 | 6.1 | 23.2 | 7.0 | 24.3 | 7.4 | 0.28 | 0.15 | 0.08 | 0.12 |
| <20 | 795 | (42.3%) | 355 | (38.4%) | 3,904 | (26.4%) |  |  |  |  |
| 20-<30 | 846 | (45.0%) | 389 | (42.1%) | 7,753 | (52.4%) |  |  |  |  |
| 30-<35 | 141 | (7.5%) | 117 | (12.7%) | 1,846 | (12.5%) | 0.36 | 0.27 | 0.07 | 0.05 |
| ≥35 | 98 | (5.2%) | 63 | (6.8%) | 1,307 | (8.8%) |  |  |  |  |
| Lactate dehydrogenase, U/L | 262.0 | 193.9 | 254.8 | 127.8 | 278.0 | 220.0 | 0.07 | 0.11 | 0.02 | 0.00 |
| C-reactive protein, mg/L | 46.3 | 50.3 | 44.2 | 49.9 | 71.8 | 67.6 | 0.39 | 0.41 | 0.06 | 0.05 |
| Lymphocyte, ×10<sup>9</sup>/L | 1.2 | 3.2 | 1.2 | 2.1 | 1.1 | 1.3 | 0.06 | 0.05 | 0.00 | 0.04 |

Notes: NA = not applicable; SD = standard deviation; SMD = standardized mean difference
\* Fully vaccinated patients were defined as those with at least 2 doses of Comirnaty or 3 doses of CoronaVac.
† Age, Charlson Comorbidity Index, time from symptom onset to hospitalization, and laboratory parameters at admission are presented in mean ± SD.

The crude incidence rate of all-cause mortality was 19.98 events per 10,000 person-days among molnupiravir users (Table 2), and 10.28 events per 10,000 person-days among nirmatrelvir/ritonavir users (Table 3). Oral antiviral use was associated with significantly lower risks of all-cause mortality (molnupiravir: HR=0.48, 95%CI=0.40-0.59, p<0.0001; nirmatrelvir/ritonavir: HR=0.34, 95%CI=0.23-0.50, p<0.0001) and the composite outcome of disease progression (molnupiravir: HR=0.60, 95%CI=0.52-0.69, p<0.0001; nirmatrelvir/ritonavir: HR=0.57, 95%CI=0.45-0.72, p<0.0001) than non-use, which was consistently observed for a reduced need for oxygen therapy (molnupiravir: HR=0.69, 95%CI=0.57-0.83, p=0.00011; nirmatrelvir/ritonavir: HR=0.73, 95%CI=0.54-0.97, p=0.032) (Tables 2 and 3, and Figure 2). Meanwhile, the relatively lower risks of IMV initiation observed in oral antiviral users were not significantly different from their control counterparts (molnupiravir: HR=0.42, 95%CI=0.17-1.01, p=0.052; nirmatrelvir/ritonavir: HR=0.97, 95%CI=0.31-3.03, p=0.96). Time to achieving lower viral burden was significantly shorter among oral antiviral users than matched controls (molnupiravir: HR=1.38, 95%CI=1.15-1.64, p=0.00046; nirmatrelvir/ritonavir: HR=1.38, 95%CI=1.07-1.79, p=0.013). There was a significant increase in the Ct value between baseline and day 5-7 in molnupiravir group (mean=6.67, 95%CI=5.91-7.43, p<0.0001), nirmatrelvir/ritonavir group (mean=7.25, 95%CI=5.93-8.56, p<0.0001), and control group (mean=3.93, 95%CI=3.57-4.28, p<0.0001). Molnupiravir users (diff=2.50, 95%CI=1.34-3.66, p<0.0001) and nirmatrelvir/ritonavir users (diff=2.86, 95%CI=0.96-4.76, p=0.0034) had larger increases in Ct value over the window of 5-7 days than their matched control groups, respectively. Amongst survivors, no significant differences were observed for hospital LOS between nirmatrelvir/ritonavir users (N=858) and matched controls (N=798), while a slightly shorter LOS was evident in molnupiravir users (N=1,706) compared to their control counterparts (N=1,561) (diff: –0.68 day, 95%CI: –1.31 to –0.06, p=0.033). Results of subgroup and sensitivity analyses were generally in line with those of the main analysis (Supplementary Tables 3 and 4, Appendix p.5-12, respectively), except for a seemingly lack of significant benefits among younger patients (aged ≤65 years) and those who had been fully vaccinated.

**Table 2.**
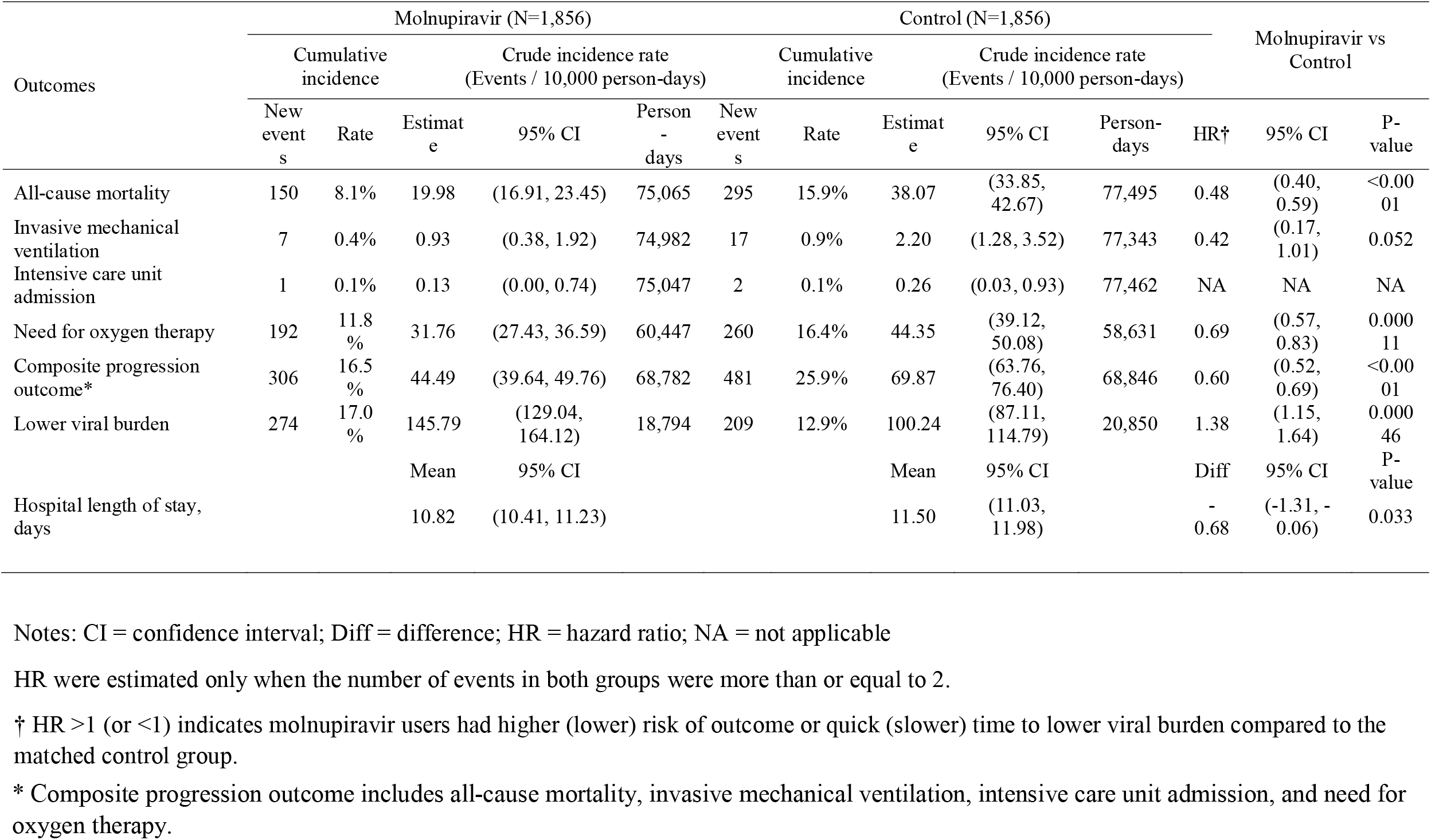
Hazard ratios of clinical and virologic outcomes for molnupiravir users versus their matched controls, and differences in hospital length of stay between the groups amongst discharged survivors

**Table 3.**
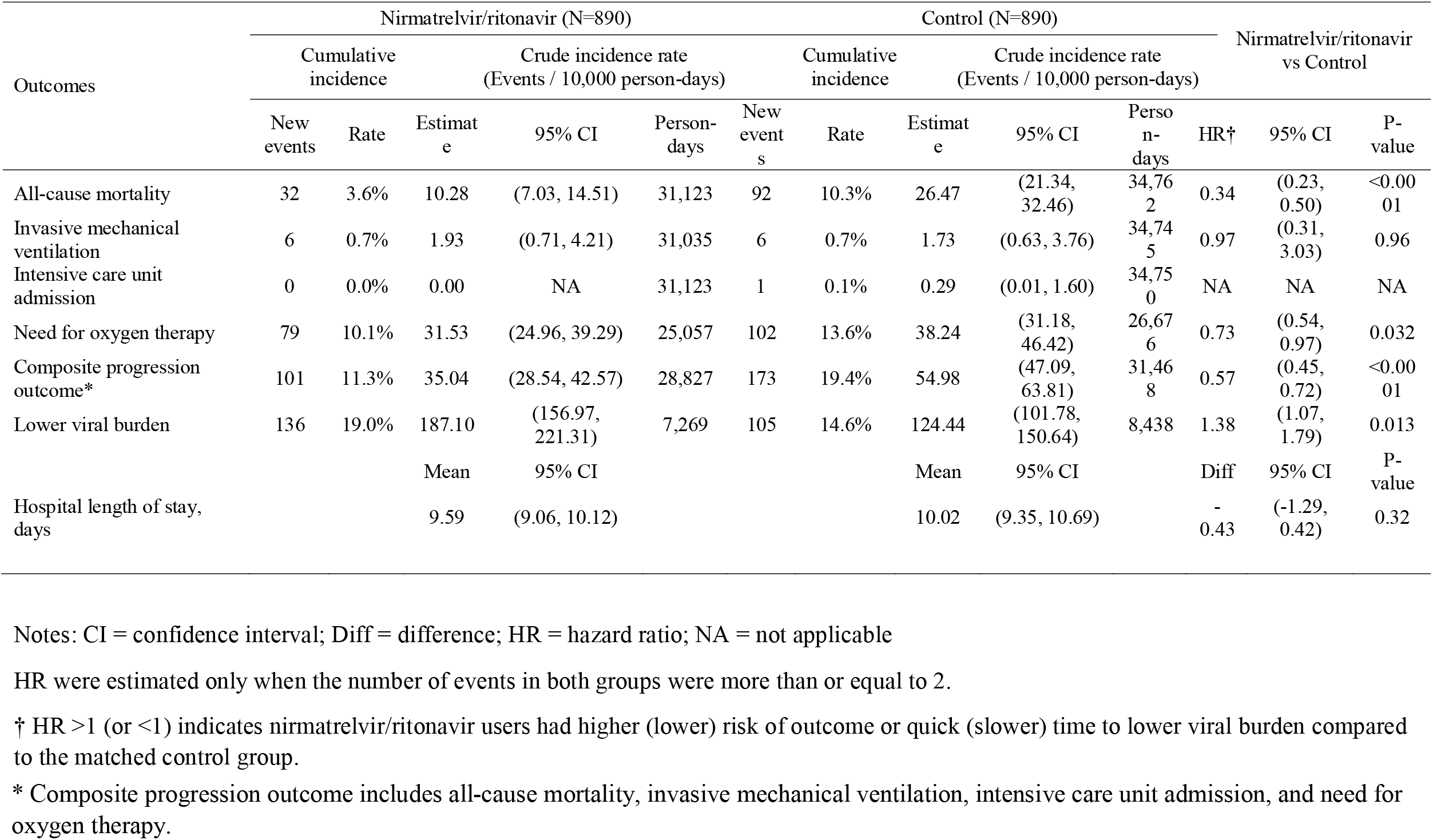
Hazard ratios of clinical and virologic outcomes, and hospital length of stay for nirmatrelvir/ritonavir users versus matched controls, and differences in hospital length of stay between the groups amongst discharged survivors

**Figure 2.**
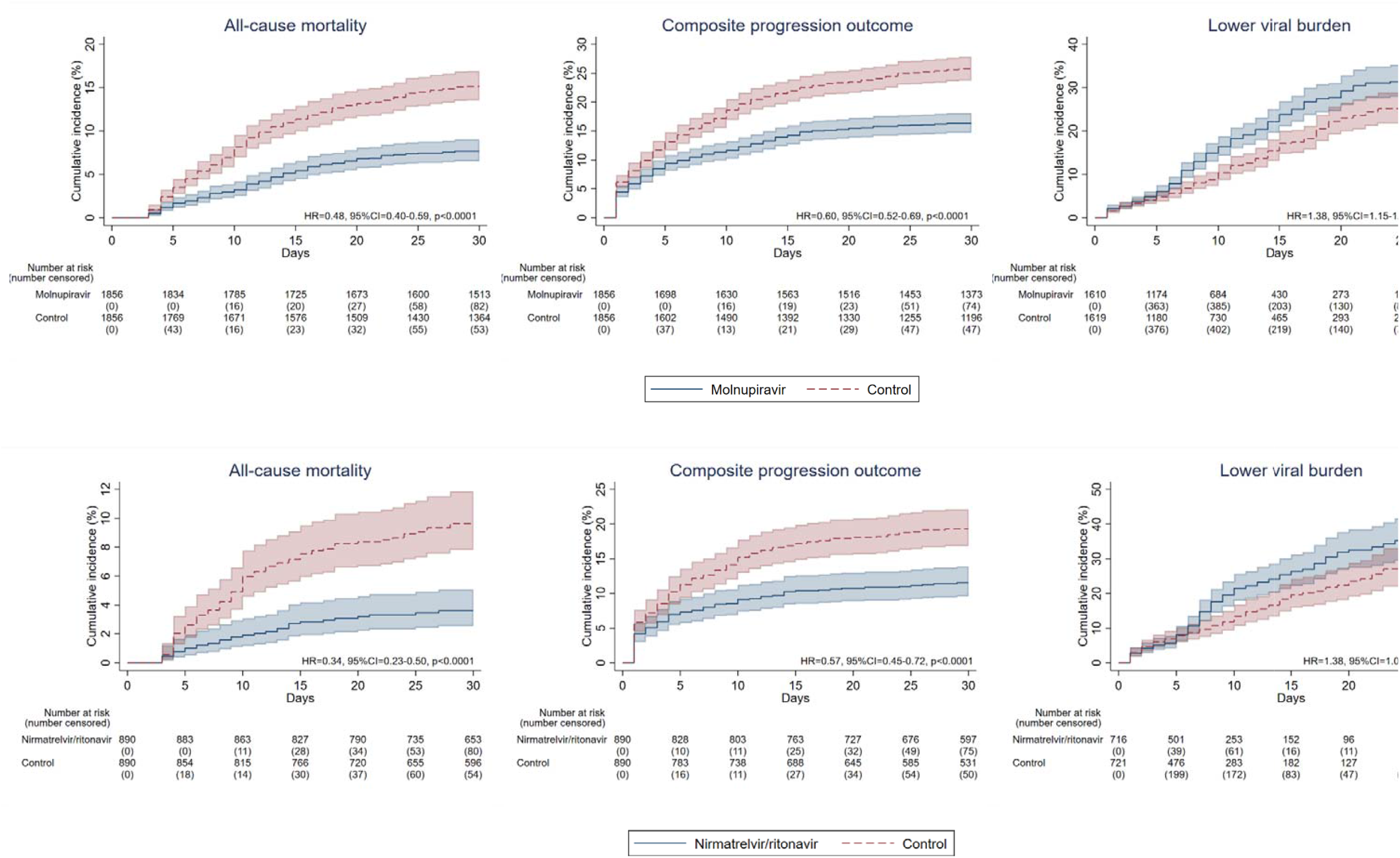
Cumulative incidence plots of (a) all-cause mortality, (b) composite progression outcome, and (c) lower viral burden for molnupiravir users versus their matched controls, and (a) all-cause mortality, (b) composite progression outcome, and (c) lower viral burden for nirmatrelvir/ritonavir users versus their matched controls

Since day-7 from baseline, the proportion of patients with in-hospital death was noticeably higher in the control group than oral antiviral users (molnupiravir: 5.3% [98 of 1,856] vs 2.3% [43 of 1,856]; nirmatrelvir/ritonavir: 3.6% [32 of 890] vs 1.3% [12 of 890]), which persisted until day-28 of follow-up (molnupiravir: 14.9% [276 of 1,856] vs 7.5% [140 of 1,856]; nirmatrelvir/ritonavir: 9.3% [83 of 890] vs 3.5% [31 of 890]) (Figure 3). At day-28, the proportion of patients discharged was higher among oral antiviral users than respective matched controls (molnupiravir: 84.4% [1,566 of 1,856] vs 75.3% [1,398 of 1,856]; nirmatrelvir/ritonavir: 89.6% [797 of 890] vs 82.5% [734 of 890]).

**Figure 3.**
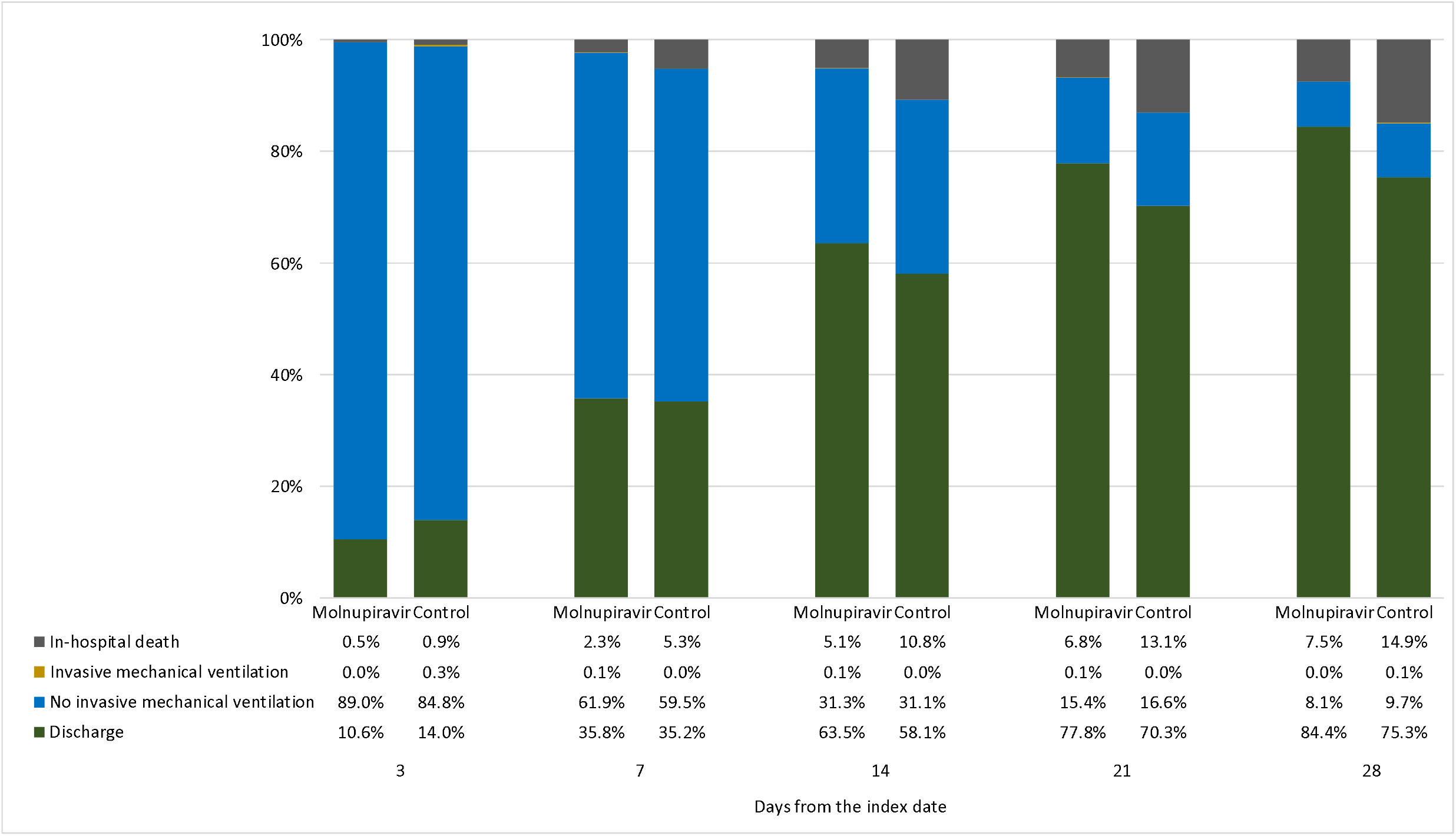
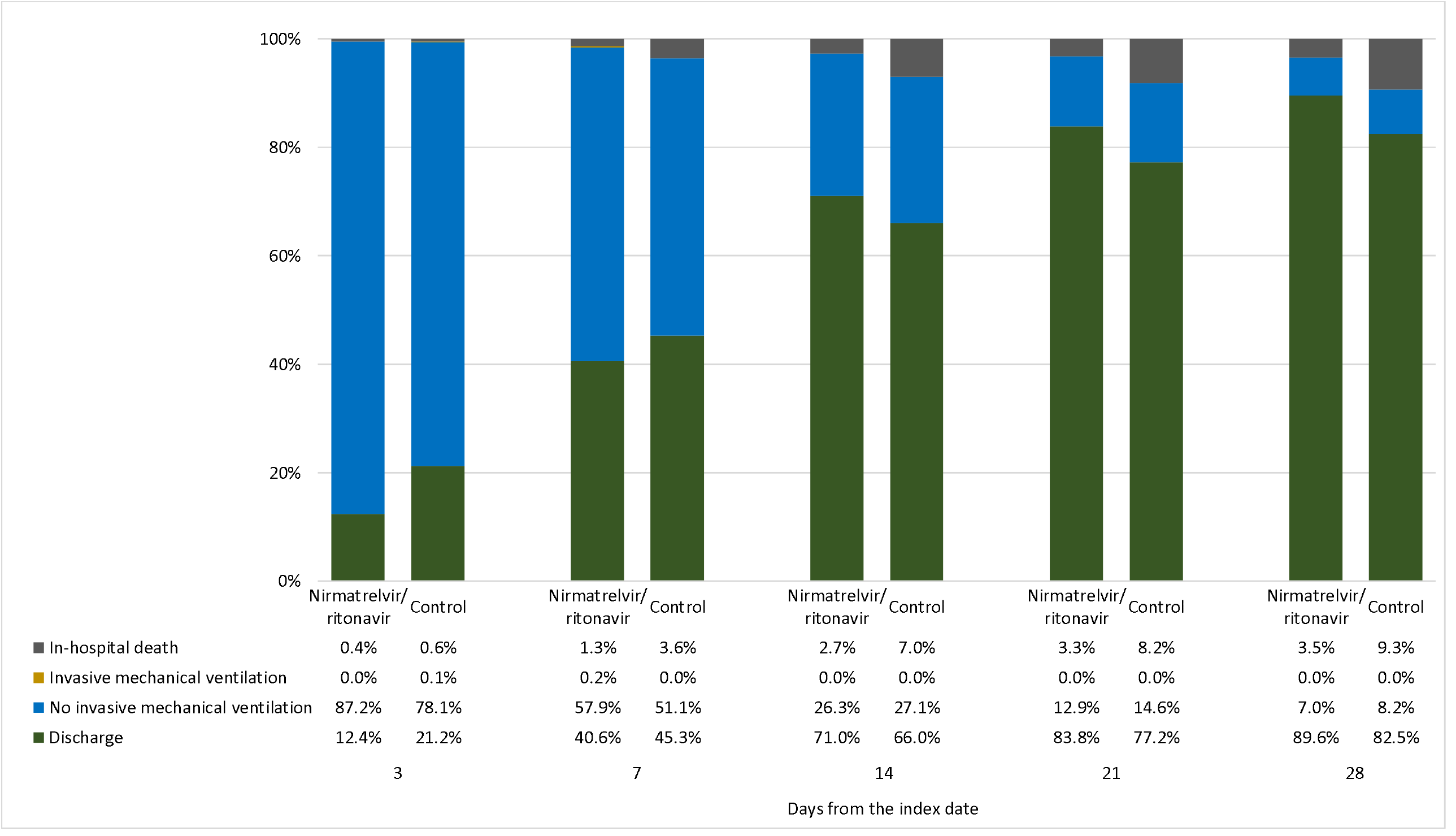
Comparison of disease status at days 3, 7, 14, 21, and 28 after the index date (hospital admission) a) between molnupiravir users and their matched controls, and b) between nirmatrelvir/ritonavir users and their matched control

## Discussion

In this retrospective cohort of COVID-19 patients not requiring any supplemental oxygen on admission, initiation of molnupiravir or nirmatrelvir/ritonavir was associated with significantly lower risks of all-cause mortality and disease progression, in addition to achieving low viral burden faster than their respective matched controls. Oral antiviral use was also associated with a reduced need for oxygen therapy. To our knowledge, this is the first real-world study exploring the inpatient use of oral antivirals during a pandemic wave dominated by the SARS-CoV-2 Omicron BA.2 subvariant.

Based on the very limited studies on the safety and efficacy of oral antivirals in COVID-19 patients, current guidelines and the medical community are now prioritizing their distribution to those who do not require supplemental oxygen, but who are at the highest risk of disease progression, i.e. who will likely benefit the most from antivirals.^4,11,25,26^ Our study cohort reflected such prescription pattern in real-world clinical practice; and provided real-world evidence supporting their use in those at risk of progression to severe disease, namely the elderly with multiple pre-existing comorbidities and who had not been fully vaccinated, during a pandemic wave dominated by the Omicron variant. The significant risk reduction in disease progression associated with both molnupiravir and nirmatrelvir/ritonavir was mainly driven by a substantial reduction in mortality risk, which has been illustrated in respective major clinical trials conducted prior to the Omicron wave (when the major circulating VOC was Delta),^27,28^ and some recent studies of nirmatrelvir/ritonavir during an Omicron surge.^29,30^ Despite an inpatient setting of the current study, our patient population who did not require any supplemental oxygen at baseline was likely different from that of the MOVe-IN trial, where the majority presented with moderate-to-severe COVID-19 and approximately half of the patients were on oxygen therapy.^31^ Also, our molnupiravir users might not be comparable to those of the MOVe-OUT trial, where the antiviral was initiated early to non-hospitalized patients with mild-to-moderate COVID-19.^28^ A secondary analysis of MOVe-OUT trial has identified a reduced need for respiratory interventions among molnupiravir users than those treated with placebo, including the patient subgroup who were hospitalized after randomization.^32^ Notably, our results established a significant mortality benefit and reduced disease progression (of increasing oxygen needs) among molnupiravir users who were hospitalized and not requiring any supplemental oxygen on admission, whilst these were not evident in the MOVe-IN trial when it was initiated at a later and more severe stage of COVID-19.^31^

In terms of viral burden reduction, our patients managed to achieve low viral burden faster with molnupiravir or nirmatrelvir/ritonavir use upon SARS-CoV-2 infection of the Omicron variant, which added clinical support to the efficacy of oral antivirals demonstrated in experimental studies.^33-38^ In recent studies based on previous VOC (including Delta), early initiation of molnupiravir has been shown to promote clinical improvement and symptom resolution in patients with mild-to-moderate COVID-19, in addition to accelerating viral burden reduction, SARS-CoV-2 RNA clearance, and elimination of infectious virus.^28,39-41^ The EPIC-HR trial was also conducted prior to the prevalence of Omicron variant, where nirmatrelvir/ritonavir use was associated with significant viral burden reduction of Delta variant in patients with mild-to-moderate COVID-19 compared to placebo.^25,27^ To the best of our knowledge, this is one of the first clinical studies offering real-world evidence of oral antiviral use on reducing viral burden among COVID-19 patients during a pandemic wave of Omicron BA.2 subvariant. This is consistent with faster viral RNA clearance identified with molnupiravir use in the latest clinical trial conducted among hospitalized patients with mild-to-moderate COVID-19 of the Omicron variant.^42^ Meanwhile, results of our subgroup analyses seemed to suggest a lack of significant benefits in younger patients and those who had been fully vaccinated, which would add support to prioritize the prescription of oral antivirals to the elderly and those with inadequate vaccination, who were also likely at a higher risk of progression to severe COVID-19. Likewise, recent studies of nirmatrelvir/ritonavir during an Omicron surge have suggested significant clinical and mortality benefits in the elderly, yet insufficient evidence for younger patients.^29,30^ Nevertheless, further research on the real-world effectiveness of oral antivirals in specific patient populations is needed, as our results could be confounded by the limited sample size, and hence the small number of events, in certain patient subgroups.

This territory-wide, retrospective cohort study of COVID-19 patients who did not initially require supplemental oxygen suggested that initiating oral antivirals within 2 days of admission was associated with significant risk reduction in all-cause mortality and disease progression, and achieving low viral burden faster compared to non-use. Referring to the medical records of hospitalized cases who were closely monitored, clinical outcomes and procedures were systematically documented and analyzed. Medication adherence could also be guaranteed in an inpatient setting compared to oral antiviral users in the community. Nevertheless, several limitations of our study should be acknowledged. Firstly, we cannot exclude the possibility of selection bias or confounding by indication in this observational study, despite our population-based cohort was fully representative of the local COVID-19 patient population who did not require supplemental oxygen on admission. Besides, the clinical profile of our patients who would be deemed at risk of progression to severe COVID-19 might be different from those in the major trials of molnupiravir and nirmatrelvir/ritonavir, for instance, their dominant risk factor was overweight or obesity,^27,28^ whilst ours was old age. Moreover, since our study was retrospective, patients that received oral antivirals could be those considered in more need for treatment than those that remained untreated, despite balance on propensity score weighting of variables including those indicating severity. Unfortunately, information on symptom onset date in most of the patients, oxygen saturation, respiratory rate, and pulse rate that might have been appropriate indicators of illness severity were unavailable for this retrospective study. Secondly, our results could potentially be biased considering clinical contraindications by drug-drug interactions for nirmatrelvir/ritonavir, or patient preferences to avoid molnupiravir due to concerns about possible mutagenicity on fertility or pregnancy.^43^ However, our analysis excluded patients with drug contraindications to nirmatrelvir/ritonavir, and those with severe renal or liver diseases for fair comparisons between oral antiviral users and matched controls. Thirdly, since the Ct value was no longer adopted as one of the discharge criteria during our study period, patients might have already been deemed clinically stable for discharge before reaching the specific cutoff. Furthermore, the interpretation of our viral burden results could be dependent on the efficiency of sampling, specimen type, and limited by the lack of clinical data on viral infectiousness. While all hospitals shared the same standard care protocol for COVID-19 patients, including discharge criteria, there was no clear and consistent documentation in the electronic health records. As such, we caution that our LOS outcome might be specific and not generalizable to other settings. Accordingly, further studies are needed to confirm our findings on viral burden reduction and LOS associated with oral antiviral use. Lastly, the generalizability of our findings could be undermined by an inpatient setting of our cohort, and some of our subgroup analyses were likely underpowered due to their small sample sizes (namely younger patients and those who were fully vaccinated). Results from ongoing trials (namely PANORAMIC^44^ and RECOVERY^45^, NCT04746183, NCT05195060, and NCT05011513) and observational studies are awaiting, and further research is needed to explore the safety and effectiveness of oral antivirals in different patient populations (especially by COVID-19 vaccination status and VOC), drug combinations, and other healthcare settings such as nursing homes or residential care facilities.

As proposed by the medical and research community, logistics and distribution issues should be adequately addressed by governments and the healthcare sector to meet ethical standards and promote optimal and equitable access in the face of limited supplies, such as developing an evidence-based scoring system or risk prediction tools to help physicians prioritizing the distribution of oral antivirals to COVID-19 patients who would most likely benefit from them, based on predicted efficacy and risk assessments.^11,25,26^ Notably, some unknown long-term risks associated with molnupiravir use include possible carcinogenicity and teratogenicity, as mutations have been observed in mammalian cells *in vitro*; and the risk of emergence of more infectious and vaccine-resistant viral variants attributed to the genetic mutations induced.^7-9,46-48^ Moreover, concerns about the development of drug resistance to molnupiravir and nirmatrelvir/ritonavir have been raised, especially considering the high mutation rates of SARS-CoV-2 and potential selective pressure induced by an extensive use of antiviral monotherapy.^26,49^ Active pharmacovigilance programs and sequencing of viral mutations are essential to monitoring their long-term safety and effectiveness in different patient populations and waves of COVID-19 pandemic.^26^

In conclusion, this retrospective cohort study of hospitalized COVID-19 patients who did not initially require supplemental oxygen suggested that early initiation of oral antivirals was associated with significant risk reduction in all-cause mortality and disease progression, as well as achieving low viral burden faster than non-use, during an epidemic of the SARS-CoV-2 Omicron BA.2 subvariant. As both oral antivirals are currently indicated for non-hospitalized COVID-19 patients who are at high risk of disease progression, ongoing research will inform the safety and effectiveness of oral antivirals in specific patient populations, drug combinations, and healthcare settings.

## Data Availability

The clinical outcome data were extracted from the Hospital Authority database in Hong Kong and vaccination records were extracted from the eSARS data provided by the Centre for Health Protection. The data custodians (the Hospital Authority and the Department of Health of Hong Kong SAR government) provided the underlying individual patient data to the University of Hong Kong for the purpose of performing scientific research for the study. Restrictions apply to the availability of these data, which were used under license for this study. Authors must not transmit or release the Data, in whole or in part and in whatever form or media, or any other parties or place outside Hong Kong; and fully comply with the duties under the law relating to the protection of personal data including those under the Personal Data (Privacy) Ordinance and its principles in all aspects.

## Contributors

The study was designed by CKHW, GML and BJC. CKHW, ICHA, EHYL and BJC had access to the underlying data of the study. The underlying data were verified by CKHW, ICHA and EHYL. Data analyses were done by CKHW and ICHA. CKHW and KTKL wrote the first draft of the manuscript which was revised by GML and BJC. CKHW, GML and BJC were responsible for the decision to submit for publication. All authors interpreted data, provided critical review and revision of the text and approved the final version of the manuscript.

## Declaration of interests

BJC reports honoraria from AstraZeneca, Fosun Pharma, GlaxoSmithKline, Moderna, Pfizer, Roche and Sanofi Pasteur. BJC has provided scientific advice to Pfizer and AstraZeneca on issues related to disease burden and vaccine effectiveness. He has not provided scientific advice to either company related to COVID antiviral effectiveness, and he has not received any funding from Pfizer or AstraZeneca for any research on antiviral effectiveness including the current work. The authors report no other potential conflicts of interest.

## Funding

This study was supported by the Health and Medical Research Fund (reference number: COVID190210), Food and Health Bureau of the Hong Kong Government.

## Notes

### Author Declarations

Institutional review board of the University of Hong Kong / Hospital Authority Hong Kong West Cluster (reference no. UW 20-493) gave ethical approval for this work.

### Summary of Updates

In this version, we have updated the manuscript by excluding hospitalized COVID-19 patients with drug contraindications to nirmatrelvir/ritonavir, severe renal or liver impairment to further mitigate confounding by indication as much as possible, and restrict the sample to those who were as equally eligible to receive either molnupiravir or nirmatrelvir/ritonavir treatment as possible. Furthermore, we have added subgroup and sensitivity analyses which generated results that were broadly consistent with our primary analysis, and reassured the validity of our data presented for the real-world effectiveness of oral antiviral drugs. The head-to-head comparison between molnupiravir and nirmatrelvir/ritonavir was removed.

