## Supplementary Information for "Real-world effectiveness of early molnupiravir and nirmatrelvir/ritonavir among hospitalized, non-oxygen-dependent COVID-19 patients on admission during Hong Kong’s Omicron BA.2 wave: an observational study" (from another version of this preprint: version 1, 2022-05-20)

Supplementary Table 1. Data completion rate of baseline characteristics for inpatients with COVID-19 in molnupiravir, nirmatrelvir-ritonavir and control groups before propensity score matching

| Baseline characteristics | Molnupiravir (n=2,359) | | Nirmatrelvir-ritonavir (n=1,000) | | Control (n=19,064) | |
| --- | --- | --- | --- | --- | --- | --- |
|  | N | % | N | % | N | % |
| Age | 2,359 | (100.0%) | 1,000 | (100.0%) | 21,423 | (100.0%) |
| Sex | 2,359 | (100.0%) | 1,000 | (100.0%) | 21,423 | (100.0%) |
| Pre-existing comorbidities |  |  |  |  |  |  |
| Charlson's Index | 2,359 | (100.0%) | 1,000 | (100.0%) | 21,423 | (100.0%) |
| Previous SARS-CoV-2 infection | 2,359 | (100.0%) | 1,000 | (100.0%) | 21,423 | (100.0%) |
| Fully vaccinated | 2,359 | (100.0%) | 1,000 | (100.0%) | 21,423 | (100.0%) |
| Concomitant treatments initiated at admission |  |  |  |  |  |  |
| Antibiotics | 2,359 | (100.0%) | 1,000 | (100.0%) | 21,423 | (100.0%) |
| Immunomodulators | 2,359 | (100.0%) | 1,000 | (100.0%) | 21,423 | (100.0%) |
| Dexamethasone | 2,359 | (100.0%) | 1,000 | (100.0%) | 21,423 | (100.0%) |
| Other systemic steroid | 2,359 | (100.0%) | 1,000 | (100.0%) | 21,423 | (100.0%) |
| Interferon-β-1b | 2,359 | (100.0%) | 1,000 | (100.0%) | 21,423 | (100.0%) |
| Baricitinib | 2,359 | (100.0%) | 1,000 | (100.0%) | 21,423 | (100.0%) |
| Tocilizumab | 2,359 | (100.0%) | 1,000 | (100.0%) | 21,423 | (100.0%) |
| Laboratory parameters at admission |  |  |  |  |  |  |
| Cycle threshold value | 1,911 | (81.0%) | 885 | (88.5%) | 15,542 | (72.5%) |
| Lactate dehydrogenase | 1,075 | (45.6%) | 441 | (44.1%) | 9,620 | (44.9%) |
| C-reactive protein | 1,551 | (65.7%) | 560 | (56.0%) | 12,326 | (57.5%) |
| Lymphocyte | 1,744 | (73.9%) | 615 | (61.5%) | 14,653 | (68.4%) |

Supplementary Table 2. Baseline characteristics of inpatients with COVID-19 in (a) molnupiravir and respective matched control groups, and (b) nirmatrelvir-ritonavir and respective matched control groups before 1:4 propensity score matching

| Baseline characteristics | Before 1:4 propensity score matching | | | | | | | | | |
| --- | --- | --- | --- | --- | --- | --- | --- | --- | --- | --- |
|  | Molnupiravir (n=2,359) | | Control (n=19,064) | | SMD | Nirmatrelvir-ritonavir (n=1,000) | | Control (n=19,064) | | SMD |
|  | N / Mean | % / SD | N / Mean | % / SD |  | N / Mean | % / SD | N / Mean | % / SD |  |
| Age, years † | 79.8 | 13.4 | 74.5 | 18.2 | 0.30 | 77.2 | 14.0 | 74.5 | 18.2 | 0.15 |
| 18-40 | 39 | (1.7%) | 1,503 | (7.9%) | 0.34 | 30 | (3.0%) | 1,503 | (7.9%) | 0.23 |
| 40-65 | 295 | (12.5%) | 3,264 | (17.1%) |  | 148 | (14.8%) | 3,264 | (17.1%) |  |
| >65 | 2,025 | (85.8%) | 14,297 | (75.0%) |  | 822 | (82.2%) | 14,297 | (75.0%) |  |
| Sex |  |  |  |  |  |  |  |  |  |  |
| Male | 1,201 | (50.9%) | 9,899 | (51.9%) | 0.02 | 508 | (50.8%) | 9,899 | (51.9%) | 0.02 |
| Female | 1,158 | (49.1%) | 9,165 | (48.1%) |  | 492 | (49.2%) | 9,165 | (48.1%) |  |
| Pre-existing comorbidities |  |  |  |  |  |  |  |  |  |  |
| Charlson's Index † | 5.9 | 2.0 | 5.2 | 2.4 | 0.29 | 5.1 | 1.7 | 5.2 | 2.4 | 0.06 |
| 1-4 | 539 | (22.9%) | 6,214 | (32.6%) | 0.22 | 337 | (33.7%) | 6,214 | (32.6%) | 0.27 |
| 5-6 | 1,048 | (44.4%) | 7,608 | (39.9%) |  | 495 | (49.5%) | 7,608 | (39.9%) |  |
| 7-14 | 772 | (32.7%) | 5,242 | (27.5%) |  | 168 | (16.8%) | 5,242 | (27.5%) |  |
| Previous SARS-CoV-2 infection | 0 | (0.0%) | 3 | (0.0%) | 0.02 | 0 | (0.0%) | 3 | (0.0%) | 0.02 |
| Full SARS-CoV-2 vaccination* | 146 | (6.2%) | 1,787 | (9.4%) | 0.12 | 102 | (10.2%) | 1,787 | (9.4%) | 0.03 |
| Concomitant treatments initiated at admission |  |  |  |  |  |  |  |  |  |  |
| Antibiotics | 269 | (11.4%) | 2,167 | (11.4%) | 0.00 | 170 | (17.0%) | 2,167 | (11.4%) | 0.16 |
| Immunomodulators | 337 | (14.3%) | 4,208 | (22.1%) | 0.20 | 122 | (12.2%) | 4,208 | (22.1%) | 0.26 |
| Dexamethasone | 307 | (13.0%) | 3,872 | (20.3%) | 0.20 | 102 | (10.2%) | 3,872 | (20.3%) | 0.28 |
| Other systemic steroid | 24 | (1.0%) | 301 | (1.6%) | 0.05 | 23 | (2.3%) | 301 | (1.6%) | 0.05 |
| Interferon-β-1b | 17 | (0.7%) | 233 | (1.2%) | 0.05 | 4 | (0.4%) | 233 | (1.2%) | 0.09 |
| Baricitinib | 2 | (0.1%) | 39 | (0.2%) | 0.03 | 8 | (0.8%) | 39 | (0.2%) | 0.08 |
| Tocilizumab | 0 | (0.0%) | 12 | (0.1%) | 0.04 | 0 | (0.0%) | 12 | (0.1%) | 0.04 |
| Laboratory parameters at admission† |  |  |  |  |  |  |  |  |  |  |
| Cycle threshold value at admission, cycle | 22.3 | 6.2 | 24.5 | 7.2 | 0.31 | 23.3 | 6.9 | 24.5 | 7.2 | 0.16 |
| <20 | 988 | (41.9%) | 4,759 | (25.0%) | 0.38 | 372 | (37.2%) | 4,759 | (25.0%) | 0.28 |
| 20-<30 | 1,065 | (45.2%) | 10,202 | (53.5%) |  | 431 | (43.1%) | 10,202 | (53.5%) |  |
| 30-<35 | 174 | (7.4%) | 2,446 | (12.8%) |  | 130 | (13.0%) | 2,446 | (12.8%) |  |
| ≥35 | 132 | (5.6%) | 1,657 | (8.7%) |  | 67 | (6.7%) | 1,657 | (8.7%) |  |
| Lactate dehydrogenase, U/L | 266.0 | 178.3 | 284.3 | 211.0 | 0.09 | 251.7 | 125.8 | 284.3 | 211.0 | 0.16 |
| C-reactive protein, mg/L | 49.7 | 52.6 | 74.3 | 68.5 | 0.37 | 43.0 | 49.5 | 74.3 | 68.5 | 0.46 |
| Lymphocyte, ×10^9^/L | 1.2 | 2.9 | 1.1 | 1.2 | 0.05 | 1.2 | 2.0 | 1.1 | 1.2 | 0.09 |

Notes: ACEI/ARB = angiotensin-converting enzyme inhibitors / angiotensin receptor blockers; NA = not applicable; NSAID = non-steroidal anti-inflammatory drugs; SD = standard deviation; SMD = standardized mean difference

* Fully vaccinated patients were defined as those with at least 2 doses of Comirnaty or 3 doses of CoronaVac.

† Age, Charlson Comorbidity Index, and laboratory parameters at admission are presented in mean ± SD.

Supplementary Figure 1. Propensity score distributions for (a) molnupiravir and respective control groups, and (b) nirmatrelvir-ritonavir and respective control groups before and after 1:4 propensity score matching

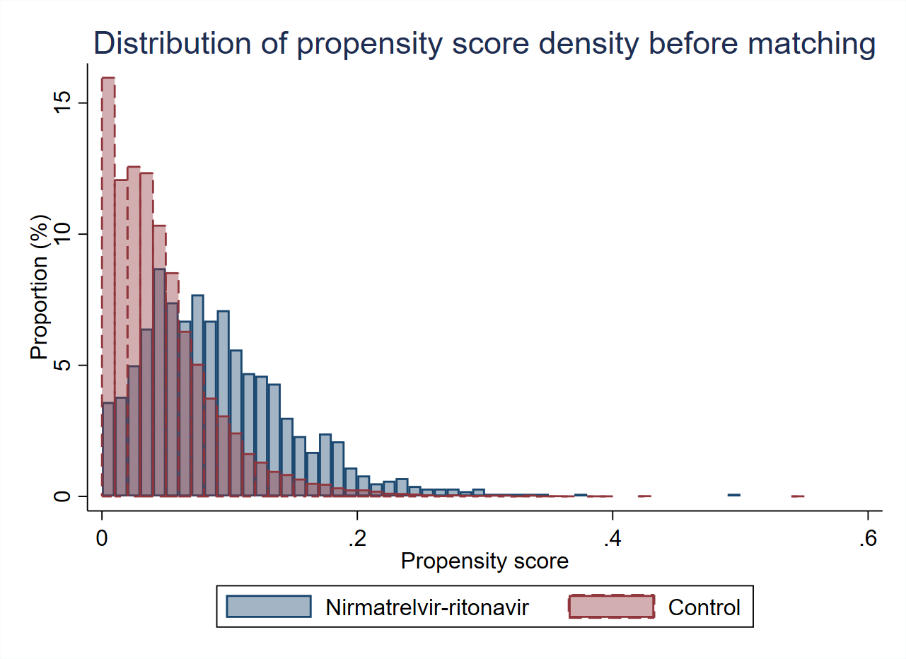

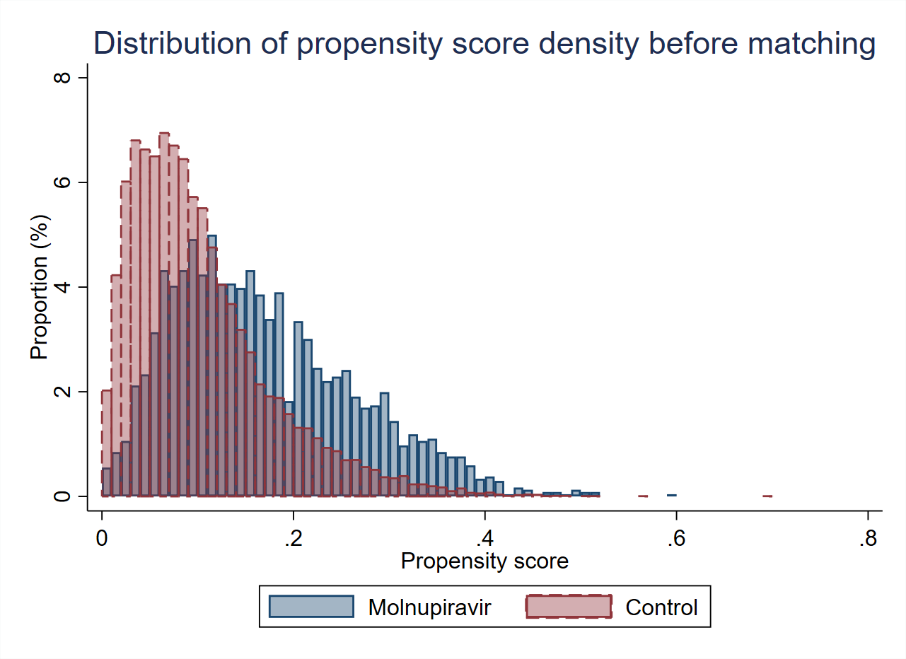

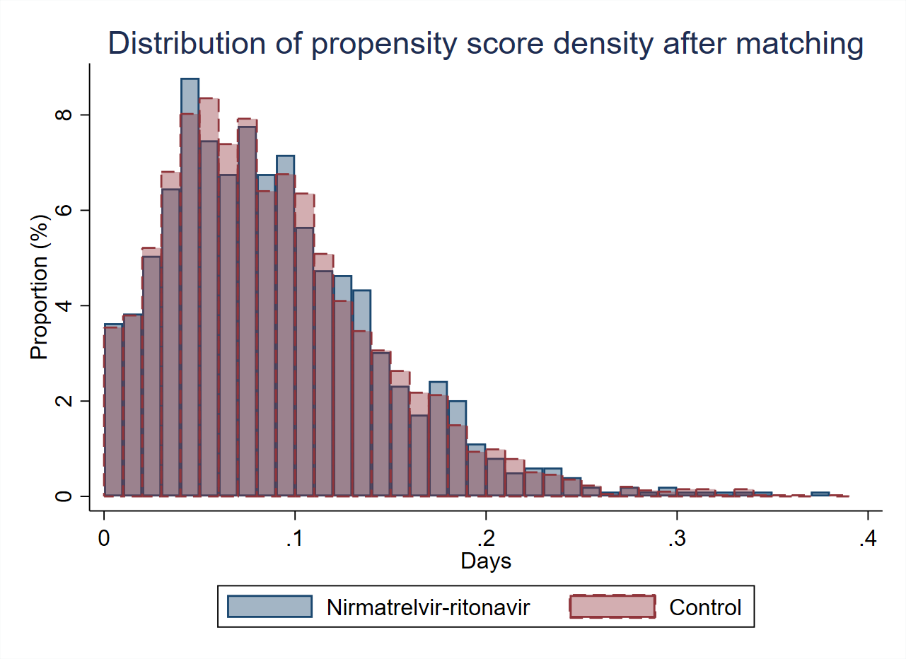

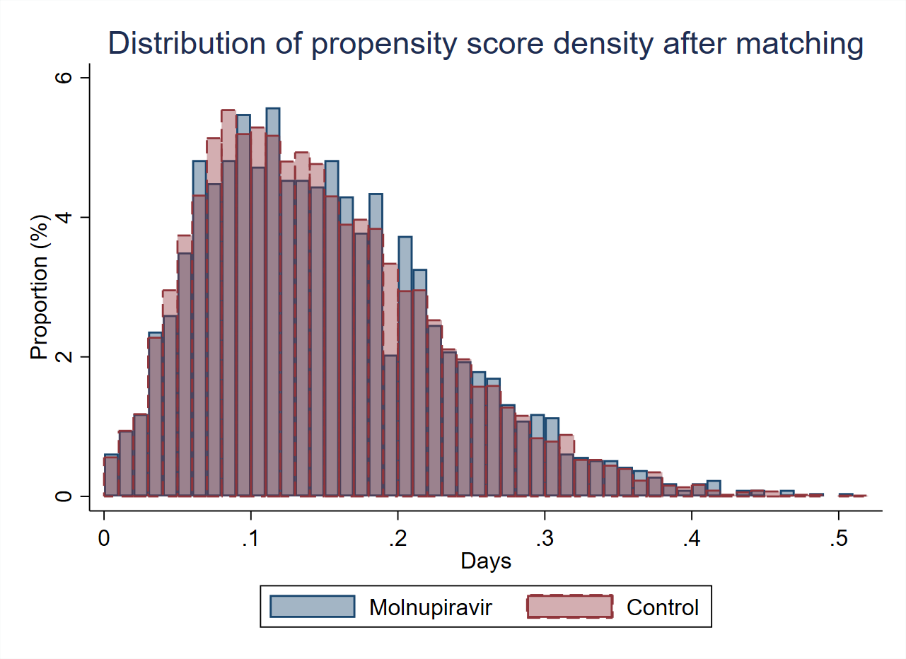

Appendix. The timeline for oral antivirals rollout in Hong Kong

| 23rd February 2022 | Introduction of new drugs for treating Coronavirus Disease 2019 | [1] |
| --- | --- | --- |
| 15th March 2022 | First shipment of COVID-19 oral drug Paxlovid distributed to HA for application | [2] |
| 3rd April 2022 | Government provides COVID-19 oral drugs to private doctors | [3] |
| 26th April 2022 | Private doctors can prescribe COVID-19 oral drugs | [4] |

References:

1. https://www.info.gov.hk/gia/general/202202/23/P2022022300303.htm

2. https://www.info.gov.hk/gia/general/202203/15/P2022031500280.htm

3. https://www.info.gov.hk/gia/general/202204/03/P2022040300028.htm

4. https://www.info.gov.hk/gia/general/202204/26/P2022042600552.htm
